# Patients with sickle cell disease presented dysregulated plasma Rb/K ratio and Gamma-glutamyl cycle in red blood cells

**DOI:** 10.1101/2023.05.17.23290113

**Authors:** Shruti Bhatt, Amit Kumar Mohapatra, Apratim Sai Rajesh, Satyabrata Meher, Pradip Kumar Panda, Ranjan Kumar Nanda, Suman Kundu

## Abstract

Patients suffering from sickle cell disease (SCD) present with multifactorial pathology, and a detailed understanding of it may help to develop novel therapeutics. In this study, the plasma elemental (^24^Mg, ^44^Ca, ^57^Fe, ^63^Cu, ^66^Zn, ^77^Se, ^85^Rb, ^208^Pb, and ^39^K) levels of SCD patients (n=10, male: 50%) and control groups (trait and healthy; n=10 each; male: 50%) were profiled using inductively coupled plasma mass spectrometry (ICP-MS). Additionally, comparative global erythrocyte metabolomics of SCD (n=5, male:100%) and healthy controls (n=5, male:100%) were carried out using liquid chromatography-mass spectrometry (LC-MS). SCD patients had higher plasma ^24^Mg, ^44^Ca, ^66^Zn, ^208^Pb, and ^39^K levels and lower levels of ^57^Fe, ^77^Se, and ^85^Rb compared to controls. These changes in elemental levels, with a decreased Rb/K ratio in the SCD group, may explain the observed frequent hemolysis and severe dehydration with oxidative stress in patients. Mass spectrometry analysis of red blood cells (RBCs of SCD (n=5) and healthy controls (n=5) identified 442 unique metabolic features which separately clustered both the study groups in principal component analysis (PCA). A set of 136 features showed differential (p<0.05; log_2_fold change>±1) regulation and was involved in D-glutamine/D-glutamate, sphingolipid, arginine biosynthesis, glutathione and glycine, serine and threonine metabolism. Interestingly, higher pyroglutamic acid levels were observed in the sickle shaped-RBCs indicating a perturbed gamma-glutamyl pathway in SCD patients. Supplementation of the depleted trace metals and targeting the perturbed metabolic pathways in the RBCs of SCD patients may provide avenues for the development of alternate therapeutics.

**Graphical abstract:** 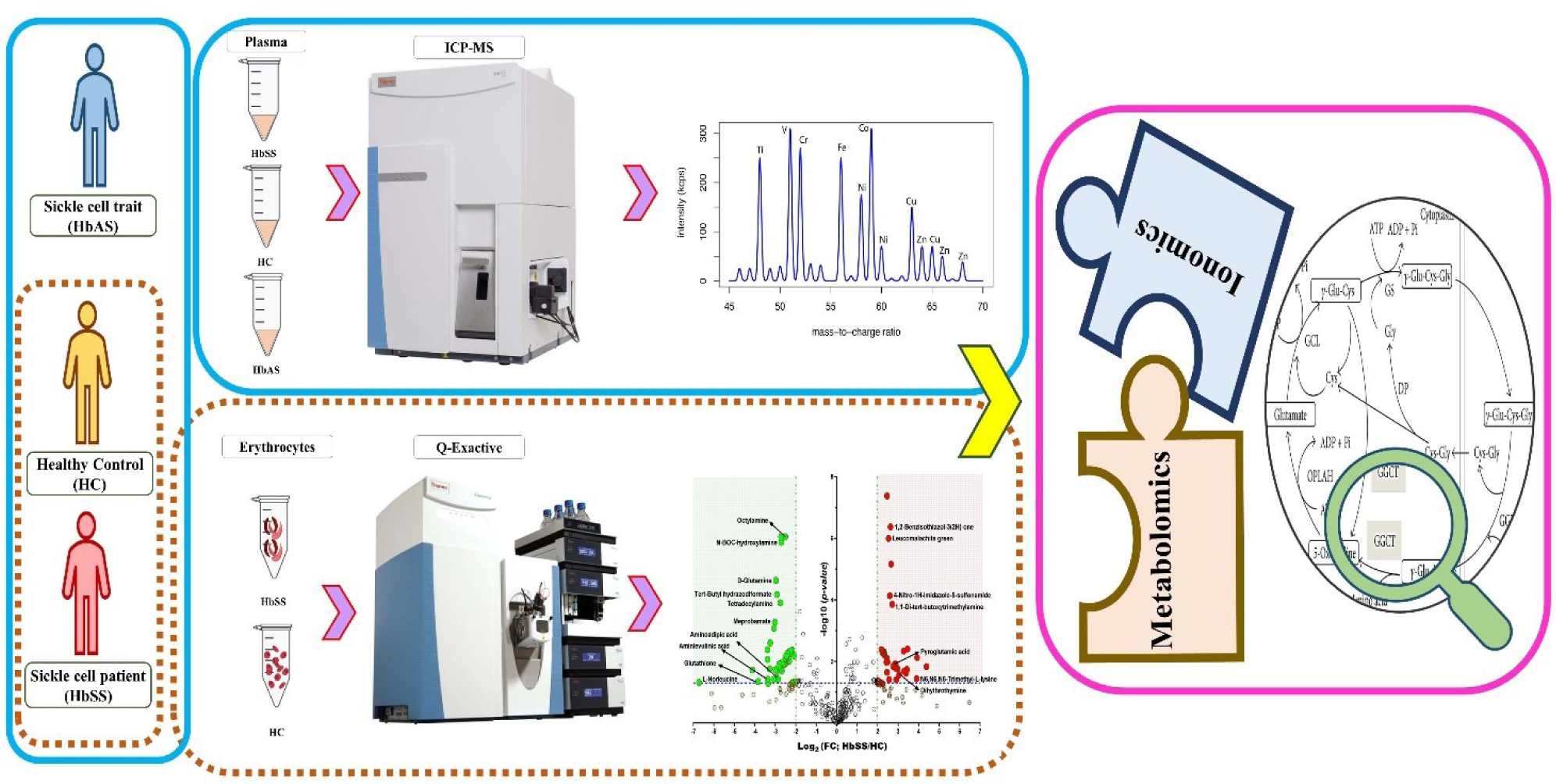

## 1. Introduction

WHO has recognized sickle cell disease (SCD), the first known human molecular disorder, as a global health pandemic. SCD cases are predicted to increase from 300,000 to 400,000 by 2050 and the majority (2/3) of it was recorded in Africa and India [1-5]. SCD is caused by a genetic mutation that translates into the substitution of valine by glutamic acid, at the 6th position in the β chain of the hemoglobin molecule leading to a structural variant known as Hemoglobin S (HbS) [6-8]. HbS has a reduced oxygen affinity, and deoxygenation near tissues leads to exposure of hydrophobic sites on individual T-state HbS molecules [9].

Exposed hydrophobic sites act as a nucleus on the HbS molecule, aggregating and forming a 14 nm fiber leading to the sickle shaped RBCs (SS-RBCs). SS-RBCs block the blood vessels leading to an impaired blood supply to organs causing recurrent episodes of acute pain (vaso-occlusive crisis: VOC) and chronic damage leading to poor survival [10-14]. Understanding the detailed pathophysiology of SCD at the molecular and elemental level will be useful to identify targets for better clinical management [15].

Multi-omics studies using liquid chromatography and inductively coupled plasma mass spectrometry (LC-MS/ICP-MS) are useful to capture the metabolites and elemental details to better understand the disease pathophysiology. Several metabolomic studies have advanced our understanding of various disease pathobiology and have become backbone for the development of alternative therapeutics [16]. Metabolite profiling deciphered that enhanced ADORA2B signaling led to increased plasma adenosine levels, contributing to multiple SCD-related complications [17]. It also captured an enhanced level of circulating sphingosine-1-phosphate (S1P), a bioactive lipid, in both SCD mice and humans [18]. The Sphk1-led S1P increase reprogrammes the metabolic balance via the release of glycolytic enzymes in the cytosol [19], towards the Embden Meyerhoff pathway (EMP) rather than the Hexose monophosphate pathway (HMP) [20]. The metabolic switching induces sickling via elevated production of 2,3-BPG (2, 3 bisphosphoglycerate) [21], signals inflammation and tissue damage via the S1PR1 (S1P receptor 1) receptor on immune cells (myeloid lineage) [22]. A dysregulated Land’s cycle, evident from increased lysophosphatidylcholine and arachidonic acid levels in SS-RBC, was reported using comparative blood metabolomic profiling of Sickle-Tg mice [23]. Interestingly, these few unbiased metabolic analyses have delivered a massive thrust to in-vitro, pharmacological, preclinical and human studies targeting the ADORA2B and SphK1–S1P–S1PR1–IL-6 signaling cascades to decipher alternative therapeutic strategies [24]. For example, PEG-ADA(adenosine deaminase deficiency), a clinically relevant drug used to treat ADA-deficient patients, reduced elevated adenosine levels, sickling, splenomegaly, multiple tissue damage, and pain [17]. Alternatively, a decrease in adenosine production can be achieved via CD73 inhibitor [e.g., adenosine 5′-(α,β-methylene) diphosphate], reduced sickling and multiple organ damage [25].

Similarly, a molecular antagonism of ADORA2B and SphK1 by PSB1115 and PF543, respectively, potently decreases sickling, inflammation and pain [26, 27]. However, only a fraction of these limited studies explored the metabolic profile of SS-RBCs derived from SCD patients [28-30]. Most have been performed on erythrocytes derived from cultured human RBCs and SCD-transgenic mice. Additionally, the previous studies fall short of capturing comprehensive pathological effects of sickle cell haplotypes including Senegal (SEN), Benin (BEN), Bantu or Central African Republic (CAR), Cameroon (CAM) and Arab-Indian (ARAB/AI) that represent the ethnic group or geographical region from which patients originated. To the best of our knowledge, no such investigation has been reported for Indian SCD diaspora. Thus, it would be interesting to investigate metabolic fingerprint of SS-RBCs (primarily Arab-Indian haplotype).

Furthermore, it has been repeatedly pressed that multi-omics can supplement and support pathophysiological understanding of various physiological stress [31]. Morphological, biochemical and metabolic changes/alterations induced by multiple pathological conditions can be correlated to the alterations in the concentration of elements in biofluids. The most notable example links RBC clearance with intracellular calcium levels [32, 33]. However, few studies establish the impact of stressors on the homeostatic loss of ions associated with SCD [34-36]. Inductively-coupled plasma mass spectrometry (ICP-MS) is a recent evolution of a decades-old analytical technique to measure elements at trace levels in biological fluids [37].

Thus, sparse and restricted reports on the perturbed metabolic pathways in RBCs and plasma elemental levels in SCD patients forged an exciting research avenue to be explored [29, 38-40]. In this study, a comparative plasma elemental composition was monitored between SCD and control (traits and healthy) groups and global RBC metabolome profiling between SCD and healthy controls was performed.This study provides an unique opportunity to integrate metabolomic and ionomic fingerprints to undertstand dysregulated metabolic pathways and identify markers associated with SCD. The lack of validated biomarkers for SCA severity represents a void in the state of knowledge of SCD that creates a critical roadblock in the design of clinical trials, the development of novel therapies and the emergence of precision medicine for SCD patients.

## 2. Materials and Methods

### 2.1 Ethics statement

All participants provided informed consent before participating in the study. This study was approved by the Institutional Ethics Committee of Sri Sri University, Cuttack, Odisha (SSCASRH/IEC/006/21) and University of Delhi South Campus, Delhi, India (UDSC/IEC/2021/Project/5.10.2021/4). The human studies reported in this study abide by the Declaration of Helsinki principles.

### 2.2 Study participants

For this study, blood samples were collected from a cohort of ten adult SCD patients with hemoglobin SS (HbSS, n=10, male 50%) disease (Supplementary Table S1), sickle cell trait individuals (hemoglobin AS: HbAS, n=10, male 50%) and ten healthy adults (controls, HC, n=10, male 50%). Written informed consent was obtained from all patients and controls. All protocols and procedures were approved by institutional ethics committees of the collaborating institutes handling patient samples.

### 2.3 Sample collection and processing

For this study, blood samples were collected from SCD patients in EDTA with hemoglobin SS disease and healthy adults (controls). Complete blood count (CBC) determined the clinical profile of participants; cellulose electrophoresis, PCR, and HPLC are presented in Supplementary Tables 1 and 2. The participants were enrolled according to the approved inclusion and exclusion criteria with consent (S.1.2). The inclusion criteria were a confirmed diagnosis of sickle cell disease by HPLC. Individuals who had received a transfusion were excluded from the study. All patients were receiving analgesic treatments. Blood samples, taken from patients with SCD, were characterized by their sickling properties [41]. Blood tubes were then centrifuged at 2000 ×g for 10 min to separate the RBCs and plasma and were aliquoted into equal volumes (0.5 ml). Plasma samples were stored in a ‒80 °C freezer until analysis. Samples were processed according to a biphasic liquid-liquid extraction (LLE) protocol. Briefly, RBC metabolism was quenched by extracting metabolites into methanol/water/chloroform solvents.

#### 2.3.1 Untargeted metabolite profiling of sickle RBCs

The 0.5 ml aliquots of human donor erythrocytes (Sri Sri University) that had been pre-treated with heparin anti-coagulant were placed in 2.0 ml microcentrifuge tubes (MCT). After adding internal standards, they were centrifuged at 1000 ×g and 4 °C for 2 min and then placed on ice while the supernatant was aspirated. The RBCs were washed twice with their resuspension in 1x phosphate-buffered saline (PBS) through centrifugation, and finally the supernatant was aspirated to leave an RBC pellet. These wash cycles remove non-erythrocytic metabolites and other compounds that may still be present outside the cells; however, it also delays quenching, and might leave residual traces of phosphate salts.

#### 2.3.2 Metabolite extraction

To the RBC pellet of each tube, 0.15 mL of ice-cold, ultrapure water (Milli-Q Millipore, Mississauga, Canada) was added to resuspend the erythrocytes. The tubes were first plunged into the dry ice for 30 seconds followed by 20 seconds of incubation in the water bath at 37 °C to quench metabolism and lyse the cells. After quenching with dry ice, 0.6 ml of methanol at 20 °C temperature was added, and the tubes were then vortexed to ensure complete mixing. The tubes were then plunged again into dry ice where 0.45 ml of chloroform was added to each tube. These tubes were vortexed briefly every 5 min for 30 min, and between each brief vortexing interval, they were placed in a cold bath. After 6 brief vortexes, the tubes were transferred to room temperature and 0.15 ml of ice-cold, ultrapure water (Milli-Q Millipore, Mississauga, Canada) was added to drive the phase separation between methanol and chloroform. The tubes were centrifuged at 1,000 ×g for 2 min at 4 °C so that a clear separation of the two phases could be observed above and below the compact disk of erythrocytes. After centrifugation, the tubes were transferred to a –20 °C freezer for an overnight incubation to allow residual chloroform to precipitate out of the aqueous methanol phase. The two liquid phases in each tube were transferred to separate 1.5 mL microcentrifuge tubes without disturbing the compact disk of erythrocytes or transferring any erythrocytes to the new tubes. The final volumes translated to a methanol/water/chloroform ratio of 4:2:3 for extraction and phase separation. The samples were then dried with speed-vac and resuspended in 0.2 mL LC mobile phase (97.9% ultra-pure water / 2% acetonitrile / 0.1% formic acid) [42].

#### 2.3.3 UPLC-Q-Exactive Plus Orbitrap MS/MS analysis

All the samples were analyzed using an UHPLC system (UHPLC Dionex UltiMate^®^ 3000, Dionex, Thermo Fisher Scientific, United States) that was controlled with Thermo Xcalibur software (Thermo Fisher Scientific, United States). The samples were separated using a Kinetex UPLC C18 column (100 × 2.1 mm, 1.9 µm; Phenomenex, Torrance, CA, United States). The mobile phase consisted of solvent A (0.1% formic acid) and solvent B (acetonitrile with 0.1% formic acid). Gradient elution was applied using the following optimized gradient program: A 35-min gradient at a flow rate of 0.3 ml/min with the following conditions was used for separation: 0–5 min, 1% B; 5–10 min, linear gradient from 1–3% B; 10–18 min, linear gradient from 3–40% B; 18–22 min, linear gradient from 40–80% B; 22–27 min, column cleaning at 95% B; and 27–35 min, re-equilibration at 1% B.

Mass spectrometry was performed on a Q-Exactive Plus™ Quadrupole-Orbitrap mass spectrometer (Thermo Fisher Scientific, United States) in positive ion mode. Data-dependent acquisition method was used for MS/MS of small molecules in the extractions. The complete MS settings were 70,000 resolution, 1e^6^ AGC (Automatic Gain Control), 100 ms max inject time, and 100–1500 m/z. The MS/MS settings were 35,000 resolution, 1e5 AGC, 100 ms max inject time, 1.0 m/z isolation window, and 30 dynamic exclusions. Three technical replicates were run for each extraction, and each technical replicate used a different HCD collision energy (25, 30, and 35, respectively).

Compound Discoverer^TM^ 3.0 (Thermo Fisher Scientific, United States) software was used to analyze the LC-MS/MS data for each extraction in positive ion mode.

Data normalization and analysis were carried out using MetaboAnalyst 5.0 (www.metaboanalyst.ca) [43]. Data exclusion was performed for metabolites with constant values across metabolites and interquartile filtering. Missing values were mean imputed, and normalization was performed. For univariate analysis, fold change and T-test values were calculated, followed by multiple testing correction based on false discovery rate (FDR). ROC-curve (receiver operating characteristic) analysis was also carried out for each metabolic feature, and 95% confidence intervals were calculated using bootstrapping with 1000 permutations. Multivariate exploratory analysis was performed using principal component analysis (PCA) and orthogonal projections to latent structures discriminant analysis (OPLS-DA). Permutation testing for OPLS-DA was applied to evaluate model stability to parameter addition. Linear support vector machine (SVM) classifiers were built to predict group class using Monte-Carlo cross-validation (MCCV) and balanced subsampling. A total of six SVMs with an increasing number of metabolites (maximum 100) were compared. Model evaluation was performed using ROC curves, and biomarker identification was achieved using the feature ranking method implemented in SVM (S.2.2, Fig. S3a, b, c).

### 2.4 Trace metal quantification using inductively coupled plasma mass spectrometry (ICP-MS)

Plasma samples of study groups were processed and subjected to ICP-MS to quantify ^24^Mg, ^44^Ca, ^57^Fe, ^63^Cu, ^66^Zn, ^77^Se, ^85^Rb, ^208^Pb, and ^39^K. Plasma samples (100 µl) were transferred to MG5 vials (Anton Paar, Graz, Austria), and HNO_3_ (250µl, 70%, #225711 Sigma Aldrich, St. Louis, Missouri, United States, with ≥99.999% trace metal basis) and H_2_O_2_ (50µl, 30%, 231 #1.07298.1000, Supelco, Inc. Bellefonte, Pennsylvania, USA) were added. Vials were sealed using sealers (#411860, Anton Paar). The vials containing the reaction mixtures were digested using a microwave digestion system (Anton Paar, Graz, Austria) ramped up to 300 W in 15 min, where it was held for 10 min. The digested samples diluted using trace metal-free water (18.2 MΩ×cm) for inductively coupled plasma mass spectrometry (ICP-MS) analysis. Digested samples were analyzed using ICP-MS (iCAP^TM^ TQ ICP-MS, Thermo Scientific, USA). Thermo Scientific Qtegra Intelligent Scientific Data Solution (ISDS) software was used for operating and controlling the instrument. The ICP-MS was calibrated using a multi-element standard mix (#92091, Sigma Aldrich, St. Louis, Missouri, United States) prepared in 1% HNO_3_ per the manufacturer’s instruction. Calibration plots were prepared at 1 ppb to 5 ppm concentrations and showed R^2^= 0.99. Digested samples were aspirated using a V-grooved MicroMist DC nebulizer and spray chamber of ICP-MS using a sample capillary (0.55 mm). Samples were then passed through a quartz torch with an injector having a diameter of 2 mm. Here, plasma ionized samples passes through sample cones, followed by a skimmer cone. The experiments were conducted in KED (Kinetic energy dissociation) mode to avoid any polyatomic ion interference. During the run, the nebulizer flow was set at 1.04 l/min with a pressure of 3.20 bar. The peristaltic pump revolved clockwise at 40 rpm. The quartz torch produced plasma and the exhaust was maintained at 0.49 mbar. Interface temperature was maintained at 29.98°C with a N_2_ flow of 14 l/min. The system’s sample and skimmer cones were made of nickel with orifices of 1 mm and 0.5 mm, respectively. Between sample runs, HNO_3_ (1%) was pumped through the nebulizer with a wash-out time of 30 sec to remove any carryover. The complete experiment was carried out with a dwell time of 0.1s. The average of three runs for each element concentration was calculated and exported for further statistical analysis. Comparative trace element levels between study groups (HbSS, HbAS and HbAA) were calculated considering the limit of detection and dilution factors.

#### 2.4.1 Statistical analysis

An unpaired t-test was performed using Graphpad prism 8 to identify group-specific variations and a p<0.05 was selected as significant.

## 3. Results

### 3.1 Clinical characteristics of the study participants

In this case and control study, a total of 30 study participants belonging to SCD as case and control (sickle cell trait and healthy controls) were used for plasma ionomic and RBC metabolomic profiling. The demographic and clinical characteristics of study participants are presented in Supplementary Tables S1 and S2.

### 3.2 Sickle cell anemia is associated with an altered ionomic profile

From the comparative plasma trace element analysis, significantly higher plasma levels of Mg (p<0.0001), Zn (p<0.0001), Ca (p<0.01), Pb (p<0.01), and K (p<0.01) were observed in SCD patients compared to healthy controls (Fig. 1a, c, d, e, h). Significantly lower plasma levels of Fe, Se (p<0.05), and Rb (p<0.01) were observed, indicating higher oxidative stress in SCD patients (Fig. 1b, g and i). However, Cu levels were similar between study groups (Fig. 1f). Overall, the subjects with sickle cell trait had comparable trace metal levels to the healthy controls except for Rb whose levels hovered in between the levels observed in SCD and healthy control (Fig. 1i). We found that there was a steady decrease in Rb/K levels in the SCD group compared to the healthy controls (Fig. 1j)

**Figure 1.**
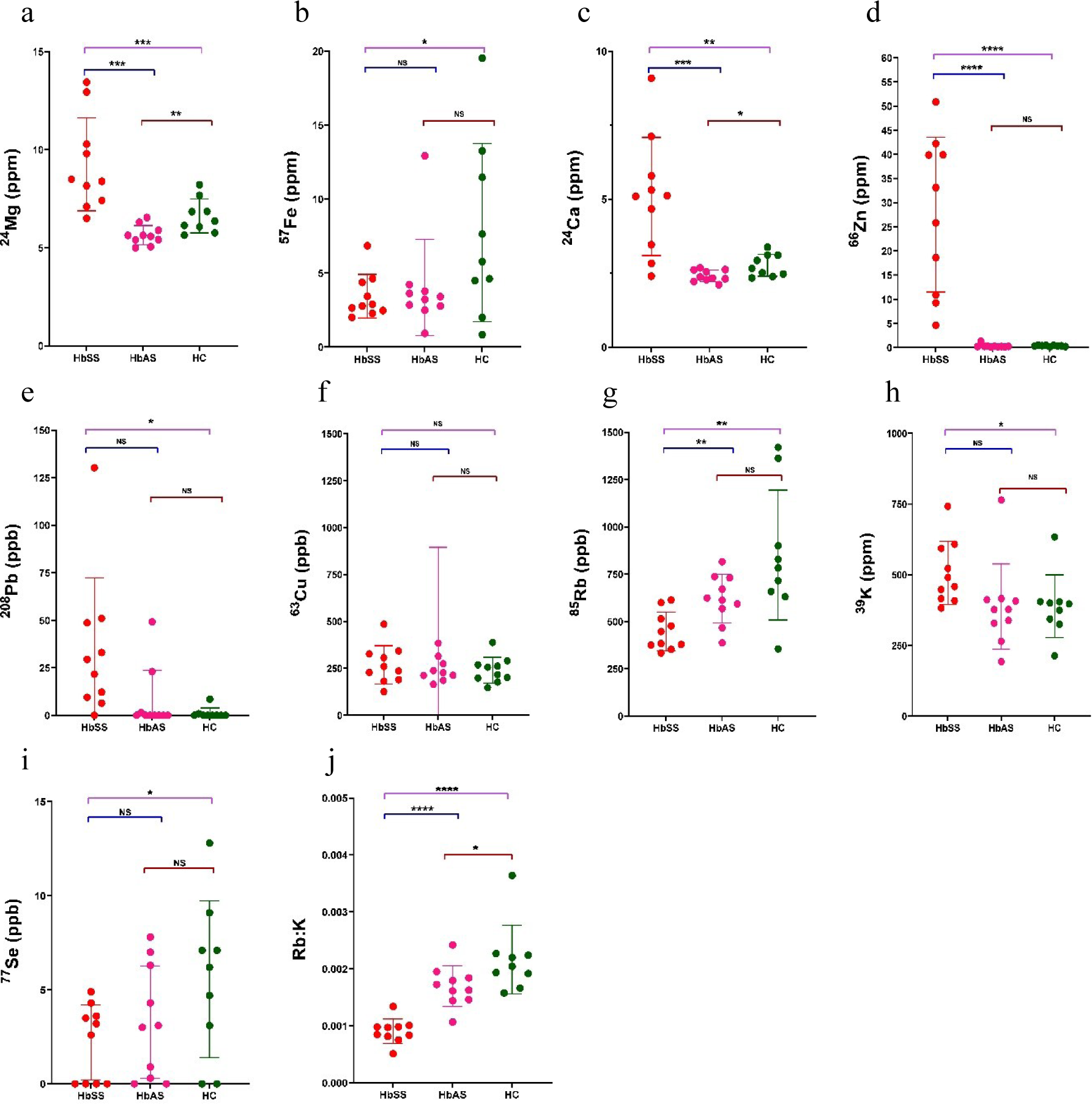
Plasma trace element levels of sickle cell patient (HbSS), sickle cell trait (HbAS) and healthy control (HC) and their ratio showed significant differences. a) Mg, b) Fe, c) Ca, d) Zn, e) Pb, f) Cu, g) Rb, h) K, i) Se, j) Rb:K. *p < 0.05, **p < 0.01, ***p < 0.001 **** p <0.0001; HbSS: Sickle cell disease; HbAS: Sickle cell trait; HC: Healthy control.

### 3.3 Metabolomics profiling of RBCs of patients with SCD demonstrated significant deregulation of specific metabolites compared to the healthy controls

Global metabolomic profiling of RBCs from SCD patients and healthy donors yielded 442 metabolite features. Principal Components Analysis (PCA) analysis of these analytes showed separate non-overlapping clusters of SCD and healthy groups. The principal components 1 and 2 explained 44.2% of the total variance (Fig. 2a). A supervised OPLS-DA model was evaluated to identify significant differentially abundant metabolites between groups and validated using permutation tests (Fig. S2a). A random permutation test (n = 2000) resulted in an interpretation rate (R_2_) and prediction ability (Q_2_) of 0.999 and 0.891, respectively (Fig. S2b). A set of 10 analytes (S.2.2, Fig. S2c) qualified the variable importance in projection (VIP) scores >1.0 and were identified as important metabolites. These metabolites displayed significantly different concentrations between groups, with fold changes >1.0 or <0.5. In the univariate analysis, 136 showed differential expression (62/56; up-/down-regulated; FC>; p<0.005) in SCD (Fig. 2b, Table S3 and Table S4). After false discovery rate adjustment, 18 analytes (7/10; up/down regulated) showed significant deregulation. In the RBC of SCD patients, glutathione, aminolevulinic acid, DL-2-aminooctanoic acid, D-glutamine, and aminoadipic acid (Table S3) levels were significantly high. Significantly lower levels of N6,N6,N6-trimethyl-L-lysine, dihydrothymine, pyroglutamic acid, 2’-alpha-mannosyl-L-tryptophan and 2-aminoisobutyric acid were observed in the RBC of the SCD patients compared to healthy controls (Fig. 2b, 2c). Hierarchical clustering analysis was performed to identify top 25 significantly dysregulated metabolic features in SCD and healthy control (Fig. 2c). The SVM model evaluation highlighted the common metabolic features (Fig. S3c) with the OPLS-DA (Fig. S2c) and hierarchical clustering analysis (Fig. 2c), i.e., thiazolidine-4-carboxylic acid, N-[(3-exo)-8-Benzyl-8-azabicyclo[3.2.1]oct-3-yl]-2-methylpropanamide, ethyl-3-phenylpropionate, meprobamate, N-BOC hydroxylamine. Interestingly, amongst the common metabolic features, thiazolidine-4-carboxylic acid and its derivatives have been reported reduce oxidative stress induced cell death [44].

**Figure 2.**
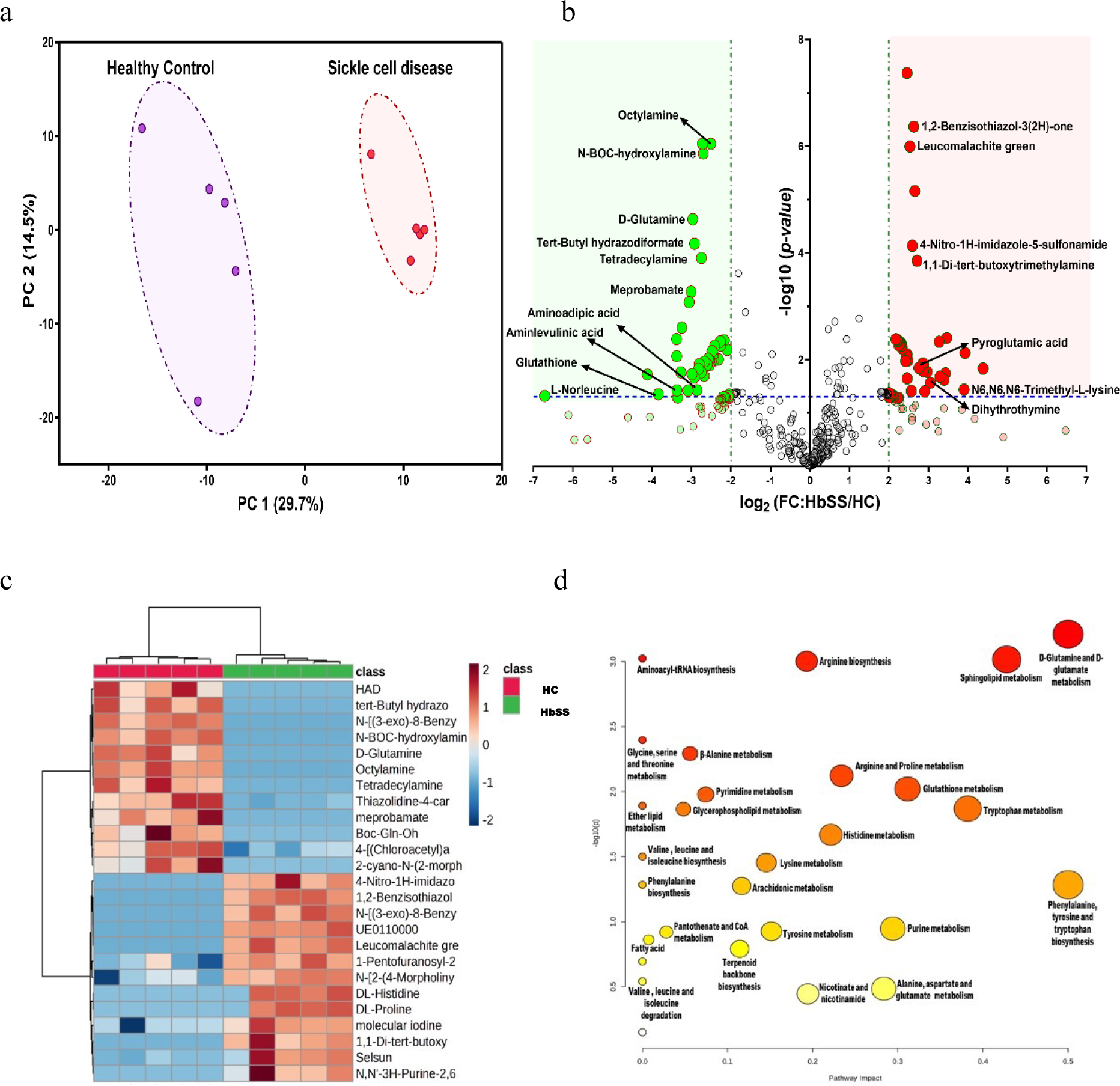
Pertubed RBC metabolic fingerprint observed in sickle cell disease patients (HbSS) compared to the healthy control (HC). a) PCA score plot showed RBC metabolites (n=442) of SCD patients cluster away from the healthy controls. b) Volcano plot highlighting the dysregulated erythrocyte metabolites in SCD (HbSS) compared to the healthy control (HC). c) Hierarchical Clustering representing the distribution of 25 important RBC metabolites in SCD patients and healthy controls. Columns correspond to samples, and rows to individual metabolites. The color scale indicates the relative abundance of metabolites: red being the most abundant and blue the less abundant metabolites. d) Metabolic pathways altered in the RBCs of SCD patients compared to healthy controls. The pathway impact values (x-axis) represent the influencing factor of topological analysis, and the – log(p) (y-axis) represents the p-value of the pathway enrichment analysis. The vital metabolic pathways were defined as having –log(p) > 2 and pathway impact factor > 0.2.

### 3.4 Functional pathway analysis

The significantly dysregulated metabolites in SCD group were selected for the KEGG pathway analysis using MetaboAnalayst (https://www.metaboanalyst.ca). D-glutamine and D-glutamate metabolism (p-value = 0.0006, Fig. S6a), aminoacyl-tRNA biosynthesis metabolism (p-value = 0.0009), sphingolipid metabolism (p-value = 0.0009), arginine biosynthesis (p-value = 0.00099), glycine, serine and threonine metabolism (p-value = 0.004, Fig. S6b), beta-alanine metabolism (p<0.005), arginine and proline metabolism (p<0.007), glutathione metabolism (p<0.009, Fig. S6c), glutathione metabolism (p<0.01), and ether lipid metabolism (p<0.012) were significantly altered in the RBC of SCD patients compared to the control group (Fig. 2d).

## 4. Discussion

The overwhelming significance of RBCs stems from the enormous abundance of hemoglobin. Any alteration in structural and functional attributes of hemoglobin can translate into irregularities in RBC functions. These irregularities can have different clinical complications inside the human body [45-47]. The RBCs are designated as a simple model to study cellular mechanisms due to less epigenetic, transcriptional and translational complications to regulate the cross-talk amongst various pathways [48-50]. However, recent omics studies highlight the existence of a complex network of cytosolic enzymes which regulate the energetics and functioning of RBCs [51, 52]. Physical and pathological stresses can alter the metabolic equilibrium and accelerate disease progression. Thus, an in-depth understanding is required to deconstruct the highly interlinked stressed RBC pathophysiology [53].

Enhanced hemolysis observed in SCD patients might be contributing to the observed higher plasma Mg level (p<0.001) [54]. This finding is in agreement with observations by Olukoga, A. O et al. (1990) [55] which establishes a negative correlation between erythrocyte and plasma Mg in the SCD cohort [56]. In contrast, low serum Mg levels have been reported in SCD patients due to Mg homeostasis [57, 58].

Similarly, RBCs contain 10-15% of the total cellular pool of calcium. Sickle cells show increased Ca as compared to normal cells in the oxygenated state [59]. Malinovská V et al. (1991) reported that stress conditions increase plasma Ca levels [60]. Higher plasma Ca levels as observed in SCD patients can be attributed to frequent hemolysis due to prolonged stress in SCD patients [61]. However, in SCD an accumulation of intraerythrocytic Ca has been reported due to malfunction of Ca^2+^ transporters [33]. Higher plasma Ca levels as observed in SCD patients can be attributed to frequent hemolysis due to prolonged stress in SCD patients [62]. An increased intracellular Ca concentration induces alteration to calcium-sensitive potassium (K^+^) channel protein 4 (also known as the putative Gardos channel and K–Cl cotransporter 1 (KCC1), KCC3 and/or KCC4 [63], resulting in potassium-efflux and decreased cell volume [64-66], which in turn increases the stiffness of RBC. Similarly, we also observed higher plasma K levels corroborating earlier reports [67-71]. Thus, K^+^ is an important indicator of SCD, however, its measurement to assess the severity in many pathophysiological conditions can be erroneous due to factors like pseudohyperkalemia and poses a challenge [72].

Another similar group I alkali metal, rubidium (Rb^+^) was significantly lower (p<0.01) in SCD patients compared to the trait and control group. It shares similar biochemical properties and readily exchanges with K^+^ and thus, can be a useful proxy for K^+^. Although its biological function still needs further understanding, ^86^Rb’s prominent presence was utilised to measure basal metabolic rate, establishing a correlation between its radioactive turnover and K^+^ concentration[73]. We are the first to report a correlation between plasma Rb and K^+^ concentration, which significantly decreased from healthy to trait and even decreased further in the SCD group. Rb may have the potential to be used as a marker to assess the severity associated with sickling. Rb has unique neurophysiological and neuroprotective properties [74-76]. Kordjazy et al. (2015) found that mice administered with Rb showed less depression-like behaviour through changes in the hippocampus [77]. Recent clinical trials and studies have widely reported that SCD patients suffer from neuro-cognitive complications [78-80]. Neurocognitive impairment have been reported in children with SCD, which affects their visuo-spatial memory(14.8%), IQ (85.4%) and copying (68.2%) [79, 81, 82]. These studies indicate that SCD can lead to the development of neuro-complications. Thus, Rb’s beneficial role in improving SCD patients’ neurocognitive complications can be further explored.

We observed significantly higher plasma lead levels in SCD compared to the trait and healthy cohort. These findings align with previous studies that have correlated BLL (Blood lead level) levels with moderate and severe anemia [83]. In addition to neurological toxicity, Pb can worsen sickle cell anemia by impairing heme synthesis and increasing the rate of red blood cell destruction [84]. Schwartz et al (1982) showed a dose-dependent increase in anemia in children with blood lead levels near 25 μg/dl [83, 85]. A linear decrease in hemoglobin was reported in children with increased BLL (BLL >30 μg/dl) by Drossos et al (1985) [86]. Although these findings indicate a relationship between Pb levels and SCD, a more detailed analysis is required to solidify these claims.

High plasma Zn levels of the SCD group was observed compared to trait and control group that does not corroborate with earlier reports [34, 87]. An increased iron (Fe) concentration in the intestinal lumen may antagonize the uptake of Zn [88]. Zn concentrations have been inversely correlated to Copper (Cu) levels [87, 89]. This depletion of Cu could impair iron absorption [90]. However, no such relationship was observed in our present study and no significant change in Cu levels was observed across the group. On the contrary, a steady Fe increase was observed in trait and healthy group (p<0.05), which indicates disrupted Fe homeostasis and most often caused by excessive urinary loss of iron as reported by some studies [91, 92].

Selenium (Se) is an essential component of mammalian enzymes like glutathione peroxidases (GPx) [93, 94], providing antioxidant defence against ROS. Erythrocytes of Se-deficient rats failed to protect hemoglobin from oxidative damage in the presence of ascorbate or H_2_O_2_ or glutathione [94]. Se deficiency is known to alter erythroid parameters like RBC, HCT, HGB, and MCHC (P < 0.05) and makes erythrocytes osmotically fragile [95]. The family is involved in oxido-reduction reactions and these reactions occur in diverse tissues and physiological pathways. Se supplementation has preventive and therapeutic role in diverse disease conditions [96-100]. Plasma Se levels were highest in the healthy and lowest in the SCD group (p<0.05); so Se supplementation may be beneficial to SCD patients.

### 4.1 Metabolic insights in sickle RBCs indicate an altered Gamma-Glutamyl cycle which fuels ATP depletion from sickle red blood cells

The main oxidative damage control system in RBCs, the glutathione pathway, has been reported to be altered in HbS red blood cells [101]. There is increasing evidence that this alteration leads to oxidative stress with a negative domino effect on the pathophysiology of SCD [61]. ROS can be derived non-enzymatically (Fenton chemistry) from denatured sickle hemoglobin (Hb S) moieties and lipid peroxidation or derived enzymatically by the action of NADPH oxidase. ROS damage RBC membranes and decrease cell deformability, which contributes to the pathophysiology of SCD [102]. Plasma-free hemoglobin (Hb) and iron chelates are by-products of hemolysis that can also act as oxidants [103]. To counteract ROS, mammalian cells have antioxidant pathways involving reduced glutathione (GSH), NAD(H), NADP(H), glutamine and nitric oxide (NO), which are complex and interlinked. Glutathione exists in a reduced (GSH) and oxidized (GSSG) form. The thiol reductant, GSH, scavenges ROS such as hydrogen peroxide and lipid peroxides [103, 104]. GSH can also interact with Hb to form glutathiol-hemoglobin (G-Hb) which reduces the propensity for sickling [105]. Similarly, we observed alterations in glutathione pathway along with disruptions in glutamine/glutamate metabolism, which are the main oxidative damages control system in RBCs of HbS cells.

Significant disruptions in glycine, serine, and threonine metabolism with lower 5-aminolevulinate levels (p-value<0.01), which is a by-product of glycine, indicate lower glycine levels (Fig. 3, Fig. S6c). Chronic oxidative stress leads to increased GSSG efflux that exceeds the rate of GSH synthesis [106]. However, rapid efflux of GSSG leads to NADPH depletion, which was observed in our study. Thus, de novo GSH synthesis becomes critical in these oxidative stress conditions and needs glycine, glutamate, and cysteine in an ATP-dependent biosynthesis. Catabolism of GSH via membrane mounted gamma-glutamyl transpeptidase (GGT) followed by removal of gamma-glutamyl moiety from GSH by gamma-glutamyl transpeptidase yields cysteinyl-glycine conjugates (Cys-Gly) and g-glutamyl-amino acid (G-Glu) as products. Hydrolysis of these conjugates by ectoprotein dipeptidases (DPT) yields cysteine and glycine. The g-Glu, glycine and cysteine enter the cell through specific transporters. The g-glutamyl with an amino acid derivative enters the cell and gamma-glutamyl cyclotransferase (γ-GCT/GGCT) converts to 5-oxoproline and the corresponding amino acid. 5-oxoprolinase (OXP) coverts 5-oxoproline (pyroglutamic acid) to glutamate in a ATP dependent manner. Oxoproline is converted to a dipeptide i.e. g-glutamylcysteine by combining glutamate and cysteine by gamma-glutamylcysteine synthetase (γ-GCS/G-GCS) in a two-step reaction which utilizes an ATP per catalytic step. This g-glutamyl cysteine can either act as substrate for GGCT to recycle to 5-oxoproline or it can be converted to GSH by the addition of glycine through GSH sythetase (GS) activity with the usage of an ATP molecule [106-108].

**Figure 3.**
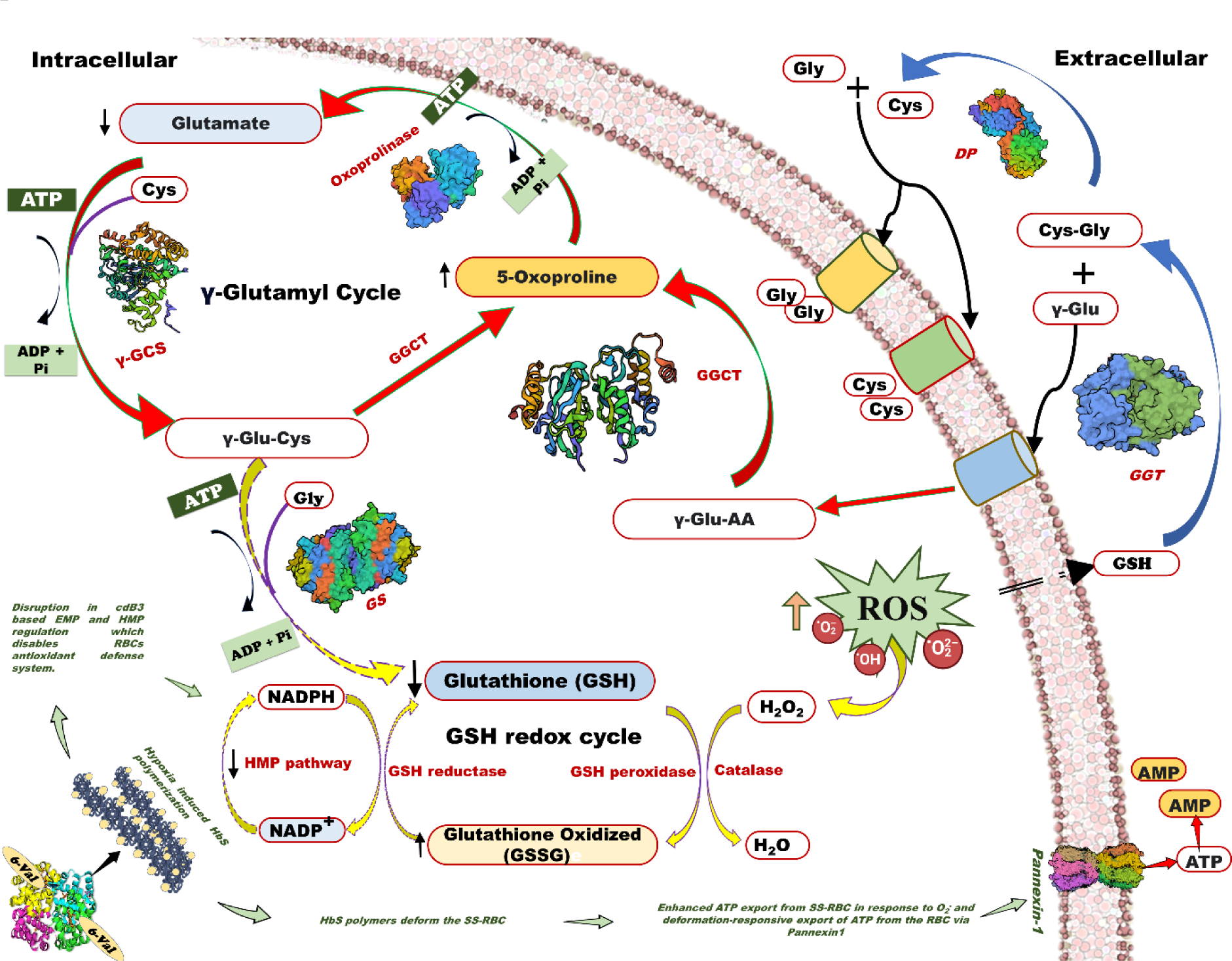
Schematics outlining the biomechanical aberration and oxidative stress leading to inoperable *γ*-glutamyl cycle in SS-RBCs. Metabolites in shades of yellow are increased in SS-RBCs compared to healthy. Metabolites in shades of blue are decreased in SS-RBCs compared to healthy. Increase in color intensity indicates a higher magnitude. The solid red arrows indicate an altered gamma-glutamyl cycle; the yellow arrow depicts altered GSH redox cycle; arrows with dashed outline entails an obstructed metabolic reaction; green arrows/text depicts previously reported pathophysiology of SS-RBCs. GGCT: gamma-glutamyl cyclotransferase; GGT: gamma-glutamyltranspeptidase; γ-GCS: glutamate cysteine ligase; GS: glutathione synthase; OXP: 5-oxoprolinase; γ-Glu-Cys-Gly: glutathione; DP: dipeptidase; HMP: hexose monophosphate pathway; EMP: Embden Meyerhof pathway; ATP: Adenosine triphosphate; ADP: Adenosine diphosphate; AMP: Adenosine monophosphate; Pi: phosphate.

ATP production and antioxidant systems within the RBC exploit Hb-based O_2_-transport to respond to various physiologic and pathophysiologic stresses. RBCs produce energy through the hexose monophosphate pathway (HMP) and glycolysis, only via the Embden–Meyerhof pathway (EMP), which generates ATP [109]. The HMP route produces NADPH, which powers the thiol-based antioxidant system critical for maintaining homeostasis in the O_2_-rich RBC [110]. For example, O_2_ offloading promotes glycolysis to generate both 2,3-BPG (a negative allosteric effector of Hb O_2_ binding) and ATP. Dynamic regulation of ATP ensures the functional activity of ion pumps, cellular flexibility, drives metabolic reactions and vaso regulation/dilation under hypoxic stress (Fig. 3). A toggle between frequency of EMP and HMP is regulated by the assembly of an EMP protein complex upon the cytoplasmic domain of the band 3 membrane protein [cdB3, also known as anion exchanger 1 (AE1)] [111-118]. Metabolite flux through EMP vs. HMP oscillates depending on the Hb conformation (oxygenation state) and cdB3 phosphorylation. RBC deoxygenation promotes the generation of ATP, while full oxygenation of RBCs promotes NADPH generation [109, 119]. Rogers and co-workers (2009) showed that RBC antioxidant systems fail when HMP flux is blunted by altered cdB3 protein assembly/phosphorylation caused by aberrant Hbs or hypoxia [109, 120, 121].

SS-RBCs are characterized by elevated indices of oxidative stress and depressed ATP levels[122], as well as elevated 2,3-DPG. Recently published evidence suggests a role for pannexin 1 (Px1) in the release of ATP from RBCs due to Gi protein stimulation in SCD [123, 124]. Zhang et al. (2011) showed that an elevated level of plasma adenosine plays a role in the increased concentration of DPG, which contributes to SCD pathophysiology by decreasing O_2_ affinity, which in successive turn promotes HbS polymerization, RBC sickling, and hemolysis [125]. As a consequence, the hydrolysis of extracellular ATP and accumulation of adenosine are favoured, and signalling via adenosine receptors may promote deoxygenation of sickle Hb and in turn (HbS) polymerization and RBC sickling.

The higher levels of pyroglutamic acid or 5-oxoproline in SS-RBCs compared to healthy counterparts can be attributed to an anomaly in salvage pathway of GSH (Fig. S6a, b). 5-oxoproline is acted upon by the 5-oxoprolinase enzyme (ATP-requiring enzyme) to yield glutamate [126]. The conversion of glutamate by the action of two consecutive ATP-dependant enzymes yields back GSH (Fig. 3). However, 5-oxoprolinase exhibited slow and inefficient enzyme activity (reaction rate of 0.45 nmol/h), which may explain the increased levels of 5-oxoproline in the cells[127]. Also, if γ-GCS fails to find an acceptor cysteine during its catalysis, it can autocyclize γ-glutamyl phosphate (intermediate product) to form 5-oxoproline [128].

Furthermore, a depleted pool of ATP, cysteine, and glycine due to sickle pathophysiology as discussed above can retard the activity γ-glutamyl cycle enzymes, i.e., OXP, γ -GCS and GS[129]. Bacchawat et al., proposed a similar futile cycle involving ATP-dependant γ-glutamyl cycle enzymes like γ-GCS and 5-oxoprolinase, leading to rapid depletion of ATP in cystinosis cells per cycle [130].

Alternatively, we can also hypothesize an abnormal activity of GGCT enzyme that leads to the accumulation of 5-oxoproline by acting on g-glutamyl-AA as a response to a decline in the concentration of cellular GSH under oxidative stress. Previous reports showed that GGCT reduces oxidative and osmotic stress in RBCs which prevents deformability prolonging their life span [131]. In various cancers, higher GGCT expression was observed and reported as a therapeutic target [132]. In SS-RBCs, a deregulated GGCT response leads to ATP depletion and also limits the availability of glycine and cysteine, which are GSH precursors [132].

## 5. Conclusion

In this study, we reported an altered elemental profile of plasma from sickle cell patients and healthy controls. We observed higher levels of Mg, Zn, Ca, K, and Pb in the plasma of SCD compared to the control groups which corroborated with frequent hemolysis, rampant dehydration of SS-RBCs via ion loss through the Gardos channel (due to K^+^ loss), and anemia-induced lead accumulation. Additionally, a steady decrease in plasma Rb/K ratio was observed for SCD when compared to trait and healthy control. We found that compromised functioning of *γ*-glutamyl cycle leading to high levels of oxidative stress was associated with SCD. These data could be validated in a larger population while taking other clinical parameters into consideration. This will be useful to gain deeper insight into the biomechanical breakdown of SS-RBCs at the molecular level and critical for identifying novel therapeutic targets for SCD patients.

## Author contributions

Shruti Bhatt: Ideated the study, processed samples, designed and performed the ionomic and metabolomic experiments, analysed and plotted data, prepared the draft of the manuscript. Amit Kumar Mohapatra: Performed ionomic experiments, analysed ionomics data, helped in drafting the manuscript Satyabrata Meher, Apratim Sai Rajesh and Pradip Kumar Panda: Recruitment of participants, clinical profiling, human sample collection, processing and transport Ranjan Kumar Nanda and Suman Kundu: Conceived and supervised the study, analysed data, provided tools and reagents, raised funds and drafted and edited the manuscript.

## Data Availability

All data produced in the present study are available upon reasonable request to the authors

## Acknowledgement

Prof. Alo Nag, Department of Biochemistry, University of Delhi South Campus, New Delhi, India, is acknowledged for administrative supervision and scientific discussion with SB. Prof. Bishnu Prasad Dash, Adjunct Professor, ICMR-Regional Medical Reasearch Centre, Bhubaneshwar, Odisha, India is acknowledged for scientific discussion with SB and ASR. Ms. Nidhi Mittal is acknowledged for scientific discussions with SB. Central Instrumentation Facility (CIF), University of Delhi South Campus, New Delhi, India, is appreciated for help with metabolomics data collection. Mr Anil Bhansali, Sai Phytoceuticals, New Delhi is appreciated for inspiring the laboratory to investigate sickle cell disease. Dharmender Singh is acknowledged for miscellaneous help to the laboratory.

## List of Abbreviation

SCD: Sickle cell disease
ICP-MS: Inductively coupled plasma mass spectrometry
LC-MS: Liquid chromatography-mass spectrometry
RBC: Red blood cell
SS-RBCs: sickle shaped RBCs (SS-RBCs)
WHO: World health organisation
VOC: Vaso-occlusive crisis
Hb: hemoglobin
HbA: adult hemoglobin
HbS: sickle cell hemoglobin
HbF: Fetal hemoglobin
VOC: Vaso-Occlusive crisis
QoL: quality of life
HSCT: hematopoietic stem cell transplantation
O_2_: Oxygen
HbS-O_2_: Oxygenated sickle hemoglobin
p50: the pressure at which hemoglobin is 50% saturated
ROS: reactive oxygen species
pO_2_: partial pressure of oxygen
Tg: Transgenic
ADORA2B: Adenosine A2b Receptor
S1P: sphingosine-1-phosphate (S1P)
EMP: Embden Meyerhoff pathway
HMP: Hexose monophosphate pathway
2: 3-DPG, 2,3 Diphosphoglyceric Acid
2: 3-BPG, 2, 3 Bisphosphoglyceric acid
S1PR1: S1P receptor 1
SphK1: Sphingosine Kinase-1
ADA: Adenosine deaminase deficiency
PEG-ADA: Polyethylene glycol-modified adenosine deaminase
CD73: ecto-5′-nucleotidase
CAR: Central African Republic (BAN or CAR)
CAM: Cameroon
BEN: Benin
SEN: Senegal
ARB/AI: Arab-Indian
HbSS: Homozygous sickle cell disease/Sickle cell disease
HbAS: Hetrozygous sickle cell trait/ Sickle cell trait
HbAA: Homozygous normal/ Healthy control
HC: Healthy control
CBC: Complete blood count
PCR: Polymerase chain reaction
HPLC: High performance liquid chromatography
HCD: Higher-energy C-trap dissociation
OPLS-DA: orthogonal projections to latent structures discriminant analysis
FDR: false discovery rate
PCA: principal component analysis
SVM: Linear support vector machine
MCCV: Monte-Carlo cross-validation
ROC: receiver operating characteristic
AUROC: Area under ROC
H_2_O_2_: Hydrogen peroxide
HNO_3_: Nitric acid
N_2_: Nitrogen
^24^Mg: Magnesium
^44^Ca: Calcium
^57^Fe: Iron
^63^Cu: Copper
^66^Zn: Zinc
^77^Se: Selenium
^85^Rb: Rubidium
^208^Pb: Lead
^39^K: Potassium
Hb-O_2_: oxyhemoglobin
KCC1/3/4: K-Cl co-transporter
IQ: Intelligent quotient,
BLL: Blood lead level
HCT: Hematocrit
HGB: Hemoglobin (g/dl)
MCHC: Mean corpuscular hemoglobin concentration
NADPH: Nicotinamide adenine dinucleotide phosphate
GSH: reduced glutathione
GSSG: oxidized (GSSG)
G-Hb: glutathiol-hemoglobin
DPT: dipeptidases (DPT)
GGCS: gamma-glutamylcysteine synthetase (G-GCS)
γ-GCT/GGCT: gamma-glutamyl cyclotransferase (G-GCT)
OXP: oxoprolinase
GGT: gamma-glutamyl transpeptidase
ATP: Adenosine triphosphate
cdB3: Band 3
γ-GCS/GGCS: gamma-glutamylcysteine synthetase
GS: GSH sythetase

