## Supplementary material for "Patients with sickle cell disease presented dysregulated plasma Rb/K ratio and Gamma-glutamyl cycle in red blood cells"

**TABLE OF CONTENTS**


| | | |
|---|---|---|
| **S.1 Supplementary methodology (extended)** | | |
| **S.1.1** | Patient recruitment | 2 |
| **S.1.2** | Informed consent | 2 |
| **S.1.3** | Study participants | 2 |
| **S.1.4** | Variables | 2 |
| **S.2 Supplementary results** | | |
| **S.2.1** | Comparative clinical profile | 3 |
| **S.2.2** | Multivariate analysis | 3 |
| **S.3 Supplementary tables** | | |
| **Table S1** | Hemoglobin profile of study participants investigated by HPLC | 5 |
| **Table S2** | Hematological parameters of participants | 6 |
| **Table S3** | Metabolites significantly upregulated in sickle cell anemia disease compared to the healthy control | 7 |
| **Table S4** | Metabolites significantly downregulated in sickle cell anemia disease compared to the healthy control. | 8 |
| **Table S5** | The important (top fourteen) metabolic pathways enriched in RBC of sickle cell disease (SCD) patients. | 9 |
| **S.4 Supplementary figures** | | |
| **Figure S1** | Comparative hematological parameters of sickle cell disease (SCD) and control (trait and healthy) study groups showed significant differences | 10 |
| **Figure S2** | RBC metabolites of sickle cell disease (HbSS: SCD) and control (HC: healthy) subjects cluster separately in OPLS-DA | 11 |
| **Figure S3** | Model performance of six SVM classifiers for sickle cell disease(HbSS: SCD) and control (HC: healthy) subjects | 12 |
| **Figure S4** | Top 25 enriched metabolic pathways observed in the RBC of SCD and healthy groups were analyzed using KEGG database | 13 |
| **Figure S5** | Identified RBC metabolites in SCD and healthy groups belong to multiple molecular classes | 14 |
| **Figure S6** | Relative abundance of the dysregulated metabolites | 15 |


.

### S.1 Supplementary methodology

**S.1.1 Patient Recruitment:** SCD patients were recruited from the sickle cell program, where they receive their medical care (Sri Sri University and Hospital, Cuttack, Odisha, India). Study team members and treating physicians recruited the study participants at the clinics and associated sickle cell infusion clinic.

**S.1.2 Informed Consent**: Potential participants (N=30) were fully informed of the purpose and activities involved in the research study during the initial contact. Interested subjects who fulfilled the eligibility criterion according to section S.1.3 were scheduled for an in-person visit where written informed consent was obtained before initiating any study procedures. Project staff conducting informed consent were appropriately trained by the Principal Investigators and had formal coursework in protecting human subjects sponsored by their institutions. Individuals with cognitive impairment or who lacked understanding were not enrolled.A copy of the signed consent was given to the patient, and other was kept in a separate locked file.

**S.1.3 Study participants**

For this study a total of 30 participants were recruited which comprised of equal number of SCD patients (n=10), sickle cell trait individuals (n=10) and healthy controls (n=10). They were recruited according to the following criterion.

a) Inclusion criterion: [i] ≥2 years of age; [ii] Homozygous sickle cell disease or S/beta 0 thalassemia SS, SC, S/β-thalassemia); [iii] Hemoglobin F ≤35%; [iv] Hemoglobin 5.5-10 g/dL; [vi] Vaso-occulusive events 1 to 10 in past 12 months; [vii] are willing to adhere to study procedures and to give written informed consent 5.1c.
b) Exclusion criteria: [i] Chronic illness and pain disorders other than SCD such as leg ulcers, fever, fractures causing pain; [ii] Transfusion within 3 weeks; [iii] unstable major psychiatric disorder such as schizophrenia or suicidal ideation; [iv] refusal to provide access to relevant medical records/Consent; [v] Participant pregnant or nursing an infant or planning pregnancy during the course of the trial.
c) Control(s) Healthy (HC) and sickle cell trait (HbAS) individuals from the same population

**S.1.4 Variables-** Demographics: Age, gender, SCD genotype and acute/chronic pain
Laboratory Studies: 1) Hematological parameters: (White Blood Cell (WBC), Red Blood Cell (RBC), hemoglobin (Hb), hematocrit (Hct), Mean Cell Volume (MCV), Mean Cell Hemoglobin (MCH), mean Cell Hemoglobin Concentration (MCHC), Red Cell Distribution Width (RDW), platelet count (Plt), and Mean Platelet Volume (MPV).

2) HPLC profiling, PCR determination of SCD

### S.2 Supplementary results

#### S.2.1 Comparative clinical profile

As part of the study, we analysed blood samples from the participants to investigate the effect of SCD on haematological parameters. We observed a significant (p-value<0.001) decrease in RBC, hemoglobin (g/dl) and haematocrit of SCD patients compared to the controls (trait and healthy). This is the evidence for predominant lysis in SCD in comparison with controls. However, a significant increase in MCHC and RDW in SCD compared to healthy controls suggests rampant dehydration of SS-RBCs and anemia. Interestingly, the haematological profile of trait had non-significant changes except for a significant increase in MCHC that could suggest that anemia and dehydration of AS-RBCs occur in trait as well (Fig. S1).

#### S.2.2 Multivariate analysis

Partial least squares-discriminant analysis (PLS-DA) (analysis revealed distinct clustering; however, due to a small sample size further analysis could lead to overfitting; thus, an Orthogonal Projections to Latent Structures Discriminant Analysis (OPLS-DA) was performed. A supervised OPLS-DA model was evaluated to identify significantly differentially abundant metabolites between groups and validated using permutation tests (Fig. S2a). A random permutation test (n = 2,000) resulted in an interpretation rate ($R^2$) and prediction ability ($Q_2$) of 0.999 and 0.902, respectively (Fig. S2b). According to the variable importance in projection (VIP) scores, the top 10 metabolites were ethyl-3-phenylpropanoic (UE0110000), octylamine, 1,2 benzisothiazol-3(2H)-one, leucomalachite, N-BOC hydroxylamine, D-glutamine, tetradecylamine, thiazolidine-4-carboxylic acid, meprobamate, tert-butyl hydrazodiformate (Fig. S2c). These metabolites displayed significantly different concentrations between groups, with fold changes >2.0 or <0.5. Support vector machine (SVM) classifiers were built to evaluate the ability of metabolic features to predict the sickle cell disease group (Fig. S3a). SVM with only five metabolites had a good AUROC (0.955, 95% CI = 0.5-1), and increasing the number of metabolites to fifteen kept the AUROC to 1 (95% CI = 1-1). Further addition of metabolites had no change in AUROC (AUROC with 100 metabolites = 1, 95% CI = 1-1; Fig. S2a). Similar observations were made for prediction accuracies (Fig. S3b), where adding ten metabolites increased accuracy by 14.5 %, compared to the model with five metabolites. However, using 100 metabolites only improved accuracy by 3 %, compared to SVM with 15 metabolites. Variables selected in the SVM model with 15 metabolites are shown in Fig. S3c. Most of these metabolites had significantly different concentrations between SCD and healthy control. The SVM model evaluation highlighted the common metabolic features with the OPLS-DA, i.e., thiazolidine-4-carboxylic acid, N-[(3-exo)-8-Benzyl-8-azabicyclo[3.2.1] oct-3-yl]-2-methylpropanamide, ethyl-3-phenylpropanoic, meprobamate, N-BOC hydroxylamine (Fig. S3c). Metabolite Set Enrichment Analysis (MSEA) was used to explore, identify and interpret the differential patterns of metabolite abundances in SCD patients and healthy controls. This approach showed that the most significantly enriched pathways were the ones related to the metabolism of sphigolipid, threonine and 2-oxo butanoate degradation, glutathione, methylhistidine, beta-alanine metabolism, pyrimidine metabolism and tryptophan metabolism (Fig. S4). Among metabolites that were significantly differentially abundant between SCD and the healthy group, organic acids, fatty acyls and benzenoids were among the top chemical classes (Fig. S5).

#### S.3 Supplementary Tables

Table S1: Hemoglobin profile of study participants investigated by HPLC

| SL NO | Genotype | PRN | HbA | $HbA_2$ | HbS | HbF |
|---|---|---|---|---|---|---|
| 1 | HbSS | 1776 | 1.10 | 3.30 | 73.20 | 21.50 |
| 2 | | 2764 | 2.90 | 2.50 | 76.10 | 17.80 |
| 3 | | 8878 | 1.70 | 2.60 | 67.60 | 28.00 |
| 4 | | 8980 | 3.10 | 3.10 | 70.60 | 22.60 |
| 5 | | 9444 | 2.60 | 3.60 | 76.10 | 16.50 |
| 6 | | 12289 | 2.80 | 2.50 | 82.10 | 12.00 |
| 7 | | 18800 | 1.10 | 3.20 | 72.20 | 22.50 |
| 8 | | 23790 | 1.50 | 1.70 | 78.80 | 17.50 |
| 9 | | 24322 | 1.20 | 1.70 | 78.90 | 17.40 |
| 10 | | 24399 | 1.80 | 3.00 | 69.80 | 22.30 |
| 11 | HbAS | RW 21 | 62.1 | 3.3 | 1.2 | 26.1 |
| 12 | | RW 16 | 59.4 | 2.9 | 1.6 | 28.8 |
| 13 | | RW1 | 63 | 3.1 | 2 | 23.9 |
| 14 | | RW36 | 60.6 | 3 | 7.5 | 23 |
| 15 | | RW22 | 63.6 | 2.6 | 1.3 | 26.1 |
| 16 | | RW 18 | 55.5 | 3.3 | 0.4 | 34.5 |
| 17 | | RW 25 | 61.2 | 3.1 | 2 | 26.4 |
| 18 | | RW10 | 64.8 | 3 | 0.7 | 24.2 |
| 19 | | RW34 | 63.9 | 3.5 | 0.5 | 24.1 |
| 20 | | RW30 | 55.8 | 2.8 | 0.7 | 34.8 |
| 21 | HbAA | RW28 | 86.9 | 2.5 | | 0.3 |
| 22 | | RW14 | 84.6 | 2.9 | | 0.5 |
| 23 | | RW18 | 86.5 | 2.7 | | 0.3 |
| 24 | | RW43 | 87.6 | 2.8 | | 0.2 |
| 25 | | RW44 | 84.6 | 2.3 | | 0.7 |
| 26 | | RW26 | 82.8 | 2.5 | | 1.4 |
| 27 | | RW31 | 84.1 | 2.8 | | 0.6 |
| 28 | | RW37 | 80.4 | 3 | | 6.2 |
| 29 | | RW8 | 85.1 | 2.6 | | 1 |
| 30 | | RW39 | 86.5 | 2.6 | | 0.5 |
| 31 | | RW7 | 87.7 | 2.7 | | 0.3 |

*HbSS: Sickle cell disease; HbAS: Sickle cell trait; HbAA: Healthy control

Table S2: Hematological parameters of participants

| Participant | Parameters | | | | | | | | |
|---|---|---|---|---|---|---|---|---|---|
| | WBC | RBC | HGB (g/dl) | HCT (%) | MCV (fL) | MCH (pg) | MCHC (g/L) | PLT ($10^3$/µL) | RDW-CV (%) |
| Sickle cell disease (SCD) | | | | | | | | | |
| SCD 1 | 11.47 | 2.97 | 9.20 | 26.8 | 90.2 | 31 | 34.3 | 316 | 14.8 |
| SCD 2 | 6 | 2.67 | 7.80 | 23.6 | 88.4 | 29.2 | 33.1 | 95 | 18.3 |
| SCD 3 | 5.42 | 1.98 | 9.80 | 26.9 | 135.9 | 49.5 | 36.4 | 337 | 14.4 |
| SCD 4 | 8.13 | 4.4 | 9.80 | 29.6 | 67.3 | 22.3 | 33.1 | 365 | 17.1 |
| SCD 5 | 10.02 | 1.95 | 4.50 | 14.1 | 72.3 | 23.1 | 31.9 | 63 | 21.3 |
| SCD 6 | 6.88 | 2.77 | 9.40 | 26.5 | 95.7 | 33.9 | 35.5 | 213 | 23.9 |
| SCD 7 | 6.23 | 3.06 | 6.90 | 20.9 | 68.3 | 22.5 | 33 | 196 | 18.6 |
| SCD 8 | 13.83 | 1.97 | 7.20 | 19.2 | 97.5 | 36.5 | 37.5 | 484 | 15.2 |
| SCD 9 | 10.18 | 3.9 | 9.80 | 30.3 | 77.7 | 25.2 | 32.2 | 222 | 18 |
| SCD 10 | 13.06 | 2.44 | 7.30 | 21.5 | 88.1 | 29.9 | 34 | 458 | 22.5 |
| Sickle cell trait (SCT) | | | | | | | | | |
| SCT 1 | 9.2 | 7.98 | 21.1 | 73.2 | 91.7 | 26.4 | 28.8 | 181 | 15 |
| SCT 2 | 7.6 | 5.23 | 12.7 | 44.5 | 85.1 | 24.3 | 28.5 | 302 | 14.1 |
| SCT 3 | 7.7 | 4.07 | 12.4 | 39.1 | 96.1 | 30.5 | 31.7 | 188 | 14.4 |
| SCT 4 | 5.8 | 3.59 | 10.7 | 33.7 | 93.9 | 29.8 | 31.8 | 238 | 15.2 |
| SCT 5 | 6.5 | 4.18 | 12.3 | 40.4 | 96.7 | 29.4 | 30.4 | 209 | 15 |
| SCT 6 | 8.1 | 4.5 | 14.3 | 45.2 | 100.4 | 31.8 | 31.6 | 250 | 18.3 |
| SCT 7 | 6.4 | 4 | 11.5 | 36.5 | 91.3 | 28.8 | 31.5 | 192 | 14.7 |
| SCT 8 | 8.9 | 5.58 | 12.6 | 40 | 71.7 | 22.6 | 31.5 | 310 | 16.7 |
| SCT 9 | 11.2 | 4.42 | 11 | 35.8 | 81 | 24.9 | 30.7 | 297 | 14.4 |
| SCT 10 | 8.3 | 4.92 | 11.7 | 37.8 | 76.8 | 23.8 | 31 | 211 | 14.4 |
| Healthy controls (HC) | | | | | | | | | |
| HC 1 | 7.6 | 3.3 | 8.3 | 29.8 | 90.3 | 25.2 | 27.9 | 275 | 13.2 |
| HC 2 | 10.7 | 6.01 | 15.6 | 53.8 | 89.5 | 26 | 29 | 313 | 14.3 |
| HC 3 | 7.4 | 3.32 | 11.2 | 35.7 | 107.5 | 33.7 | 31.4 | 206 | 12.9 |
| HC 4 | 7 | 4.53 | 11.5 | 41.6 | 91.8 | 25.4 | 27.6 | 213 | 12.3 |
| HC 5 | 6.1 | 3.62 | 8.1 | 29.6 | 81.8 | 22.4 | 27.4 | 194 | 13.4 |
| HC 6 | 7 | 5.45 | 14 | 47.7 | 87.5 | 25.7 | 29.4 | 207 | 13.2 |
| HC 7 | 9.3 | 4.81 | 10 | 43.3 | 90 | 20.8 | 23.1 | 214 | 14.4 |
| HC 8 | 5.4 | 4.78 | 14.3 | 47.8 | 100 | 29.9 | 29.9 | 156 | 17.3 |
| HC 9 | 9.4 | 4.87 | 11.6 | 42.3 | 86.9 | 23.8 | 27.4 | 220 | 13.3 |
| HC 10 | 7.7 | 5.16 | 12 | 43.9 | 85.1 | 23.3 | 27.3 | 280 | 14 |

Table S3: Metabolites significantly upregulated in sickle cell anemia disease compared to the healthy control.

| | Metabolites | $log_2(FC)$ | p-value |
|---|---|---|---|
| 1. | L-Norleucine | 6.7212 | 0.048 |
| 2. | N2-neopentyl-5-methyl-1,3,4-thiadiazol-2-amine | 4.1182 | 0.019 |
| 3. | Glutathione | 3.8398 | 0.044 |
| 4. | 2-cyano-N-(2-morpholinoethyl)acetamide | 3.38 | 0.004 |
| 5. | 3-Nitro-4-{[(tetrahydro-2H-pyran-4-yl)methyl]amino}benzenesulfonamide | 3.3785 | 0.008 |
| 6. | Aminolaevulinic acid | 3.3711 | 0.037 |
| 7. | 4-Amino-7-(1-piperidinyl)pyrazolo[5,1-c][1,2,4]triazine-3,8-dicarbonitrile | 3.2722 | 0.017 |
| 8. | Boc-Gln-Oh | 3.2456 | 0.002 |
| 9. | 3-Amino-4-hydroxy-N-methylbenzenesulfonamide | 3.0881 | 0.038 |
| 10. | Meprobamate | 3.0114 | 0.0005 |
| 11. | DL-2-Aminooctanoic acid | 2.9879 | 0.018 |
| 12. | D-Glutamine | 2.9672 | $2.34\times10^{-5}$ |
| 13. | 3-(1,1-Dioxido-2,3-dihydro-3-thiophenyl)-1,1-dimethylurea | 2.935 | 0.021616 |
| 14. | tert-Butyl hydrazodiformate | 2.9242 | $6.74\times10^{-5}$ |
| 15. | Aminoadipic acid | 2.8648 | 0.037 |
| 16. | Ro 20-1724 | 2.8241 | 0.014 |
| 17. | DL-Arginine | 2.8049 | 0.012 |
| 18. | MFCD23106399 | 2.7499 | 0.017 |
| 19. | Tetradecylamine | 2.748 | 0.0001 |
| 20. | N-[(3-exo)-8-Benzyl-8-azabicyclo[3.2.1]oct-3-yl]-2-methylpropanamide.1 | 2.7224 | $8.97\times10^{-7}$ |
| 21. | N-BOC-hydroxylamine | 2.7043 | $1.37\times10^{-6}$ |
| 22. | MNA | 2.6793 | 0.020 |
| 23. | methyl 2-(acetylamino)-4-amino-4-oxobutanoate | 2.6732 | 0.010 |
| 24. | Dimethyl 1-(3-methoxy-3-oxopropyl)-1H-pyrazole-3,5-dicarboxylate | 2.583 | 0.009 |
| 25. | L-Kynurenine | 2.5562 | 0.010 |
| 26. | ACPC | 2.5494 | 0.012 |
| 27. | Thiomorpholine 3-carboxylate | 2.5382 | 0.010 |
| 28. | Octylamine | 2.5119 | $8.97\times10^{-7}$ |
| 29. | N2-Acetylornithine | 2.5061 | 0.010 |
| 30. | L-Histidine | 2.4826 | 0.006 |
| 31. | Diethylzinc | 2.4587 | 0.007 |
| 32. | N-benzylformamide | 2.4103 | 0.005 |
| 33. | 1-Isopropyl-4-(4-isopropylphenyl)-6-(2-propyn-1-ylamino)-2(1H)-quinazolinone | 2.3183 | 0.009 |
| 34. | Muscimol | 2.27 | 0.004 |
| 35. | Dicyclohexylamine | 2.2673 | 0.005 |
| 36. | biotin amide | 2.099 | 0.006 |
| 37. | D-Proline | 2.0644 | 0.049 |
| 38. | Pyroglutamic Acid | 1.9731 | 0.045 |
| 39. | Indoleacrylic acid | 1.9215 | 0.044 |
| 40. | NP-021733 | 1.9171 | 0.041 |
| 41. | N-Boc-3-Pyrrolidinone | 1.9057 | 0.045 |
| 42. | Cyclo(D-leucyl-L-leucyl-L-leucyl-L-leucyl-L-leucyl-L-leucyl) | 1.874 | 0.041 |
| 43. | L-Tyrosine | 1.8569 | 0.040 |
| 44. | 2-((4-Nitrophenoxy)methyl)oxirane | 1.849 | 0.041 |
| 45. | Thiazolidine-4-carboxylic acid | 1.8098 | 0.0002 |
| 46. | NP-015114 | 1.7082 | 0.042 |
| 47. | Methohexital | 1.7035 | 0.007 |
| 48. | 4-[(Chloroacetyl)amino]-N-cyclopentylbenzamide | 1.6354 | 0.001 |
| 49. | L-alpha-Glycerylphosphorylcholine | 1.3093 | 0.012 |
| 50. | N-(1,1-Dioxido-2,3-dihydro-3-thiophenyl)glycine | 1.1155 | 0.022 |
| 51. | 417-720-1 | 1.1081 | 0.025 |

Table S4: Metabolites significantly downregulated in sickle cell anemia disease compared to the healthy control.

| | **Metabolites** | **$log_2$(FC)** | **p-value** |
|---|---|---|---|
| 1. | trans-3-Amino-1-Boc-4-hydroxypyrrolidine | -4.3804 | 0.014 |
| 2. | 1-((((2,5-Dioxopyrrolidin-1-yl) oxy) carbonyl) oxy) ethyl isobutyrate | -3.9234 | 0.007 |
| 3. | N6, N6, N6-Trimethyl-L-lysine | -3.8975 | 0.036 |
| 4. | N, N'-3H-Purine-2,6-diyldiacetamide | -3.4578 | 0.003 |
| 5. | Diisopropylethylamine | -3.4291 | 0.018 |
| 6. | N-4-Cbz-2-piperazinecarboxylic acid | -3.3952 | 0.024 |
| 7. | methyl (2S)-2,6-bis({[(tert-butoxy) carbonyl] amino}) hexanoate | -3.2851 | 0.020 |
| 8. | 1,4-Dinitroso-2-methylpiperazine | -3.2672 | 0.004 |
| 9. | Dihydrothymine | -3.0435 | 0.027 |
| 10. | 2-Amino-4,5,6,7-tetrahydro-1-benzothiophene-3-carbonitrile | -2.9627 | 0.016 |
| 11. | 1-[(4S)-4-Amino-5-(3-pyridinylamino) pentyl]-3-nitroguanidine | -2.9014 | 0.039 |
| 12. | cyclandelate | -2.8691 | 0.016 |
| 13. | N-{4-[(4-Methyl-1-piperidinyl) sulfonyl] phenyl}-2-thiophenecarboxamide | -2.8541 | 0.011 |
| 14. | 1,1-Di-tert-butoxytrimethylamine | -2.7075 | 0.000 |
| 15. | N-[(3-exo)-8-Benzyl-8-azabicyclo [3.2.1] oct-3-yl]-2-methylpropanamide | -2.6528 | $6.9\times10^{-6}$ |
| 16. | 1,2-Benzisothiazol-3(2H)-one | -2.6259 | $4.3\times10^{-7}$ |
| 17. | 4-Nitro-1H-imidazole-5-sulfonamide | -2.5916 | $7.3\times10^{-5}$ |
| 18. | Leucomalachite green | -2.5304 | $1.0\times10^{-6}$ |
| 19. | 2'-alpha-mannosyl-L-tryptophan | -2.4796 | 0.010 |
| 20. | UE0110000 | -2.4547 | $4.2\times10^{-8}$ |
| 21. | VO1850000 | -2.4517 | 0.007 |
| 22. | 1-(2-Furyl)-2-propyn-1-one | -2.4243 | 0.010 |
| 23. | 4-Iodoanisole | -2.3201 | 0.005 |
| 24. | 4-(p-Dimethylaminostyryl)quinoline | -2.3075 | 0.004 |
| 25. | L-Threonine | -2.3 | 0.004 |
| 26. | PEG n5 | -2.2842 | 0.005 |
| 27. | Cer(d18:1/18:0) | -2.2481 | 0.005 |
| 28. | {2-[(Phenylacetyl)amino]-3-(1H-1,2,4-triazol-1-yl)propyl}phosphonic acid | -2.2325 | 0.004 |
| 29. | 1-(chloroacetyl)pyrrolidine | -2.2178 | 0.004 |
| 30. | DL-Proline | -2.2007 | 0.004 |
| 31. | DL-Histidine | -2.1747 | 0.004 |
| 32. | 2-Aminoisobutyric acid | -2.0999 | 0.047 |
| 33. | Spermine | -2.0048 | 0.042 |
| 34. | Symmetric dimethylarginine | -1.9308 | 0.044 |
| 35. | IN00458 | -1.9143 | 0.041 |
| 36. | 3,4-Dimethoxyphenylethylamine | -1.8959 | 0.044 |
| 37. | 5-Hydroxyindoleacetic acid | -1.8695 | 0.043 |
| 38. | Vanillin | -1.866 | 0.040 |
| 39. | GO7887000 | -1.8573 | 0.041 |
| 40. | DEEMM | -1.843 | 0.041 |
| 41. | 1700191 | -1.8376 | 0.040 |
| 42. | L-Lysine hydrochloride | -1.8188 | 0.040 |
| 43. | Leucine | -1.8122 | 0.049 |
| 44. | Uracil | -1.787 | 0.010 |
| 45. | Guanosine monophosphate | -1.4496 | 0.014 |
| 46. | 1-Pentofuranosyl-2,4(1H,3H)-pyrimidinedione | -1.2388 | 0.001 |

Table S5: The important (top fourteen) metabolic pathways enriched in RBC of sickle cell disease (SCD) patients.

| Pathways | Total components | Hits | Statistic Q | Expected Q | Raw p | Holm p | FDR |
|---|---|---|---|---|---|---|---|
| Sphingolipid Metabolism | 40 | 3 | 38.172 | 11.111 | 0.001 | 0.124 | 0.111 |
| Methylhistidine Metabolism | 4 | 1 | 66.28 | 11.111 | 0.004 | 0.294 | 0.111 |
| Threonine and 2-Oxobutanoate Degradation | 20 | 1 | 64.903 | 11.111 | 0.004 | 0.343 | 0.111 |
| Glutathione Metabolism | 21 | 5 | 28.964 | 11.111 | 0.006 | 0.444 | 0.111 |
| Glycine and Serine Metabolism | 59 | 6 | 30.988 | 11.111 | 0.009 | 0.623 | 0.111 |
| Pyrimidine Metabolism | 59 | 3 | 39.401 | 11.111 | 0.009 | 0.623 | 0.111 |
| Tryptophan Metabolism | 60 | 6 | 25.151 | 11.111 | 0.015 | 0.991 | 0.144 |
| Arginine and Proline Metabolism | 53 | 6 | 26.049 | 11.111 | 0.016 | 1 | 0.144 |
| Beta-Alanine Metabolism | 34 | 5 | 28.468 | 11.111 | 0.019 | 1 | 0.153 |
| Arachidonic Acid Metabolism | 69 | 5 | 25.522 | 11.111 | 0.026 | 1 | 0.187 |
| Porphyrin Metabolism | 40 | 1 | 43.537 | 11.111 | 0.037 | 1 | 0.207 |
| Glutamate Metabolism | 49 | 6 | 26.434 | 11.111 | 0.039 | 1 | 0.207 |
| Carnitine Synthesis | 22 | 3 | 30.832 | 11.111 | 0.041 | 1 | 0.207 |
| Spermidine and Spermine Biosynthesis | 18 | 1 | 42.012 | 11.111 | 0.042 | 1 | 0.207 |

### S.4 Supplementary figures

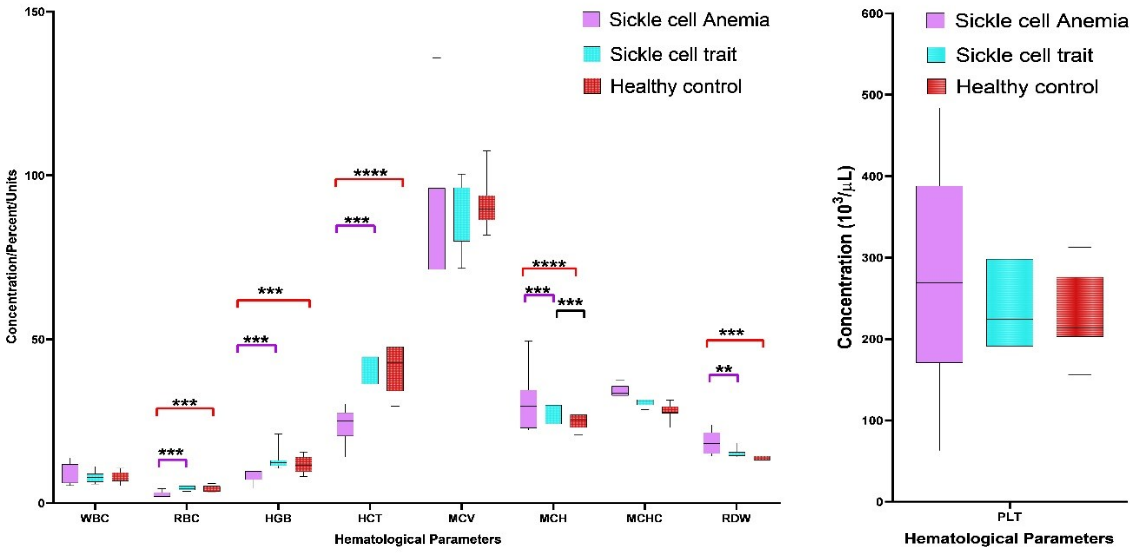


Figure S1: Comparative hematological parameters of sickle cell disease (SCD) and control (trait and healthy) study groups showed significant differences. **p < 0.05, **p < 0.01, ***p < 0.001 **** p <0.0001*; *WBC: White blood cells; RBC: red blood cells; HGB: hemoglobin (g/dl); HCT: haematocrit; MCV: Mean corpuscular volume; MCH: Mean Corpuscular Hemoglobin; MCHC: Mean corpuscular hemoglobin concentration; PLT: Platelet; RDW-CV: red cell distribution width.*

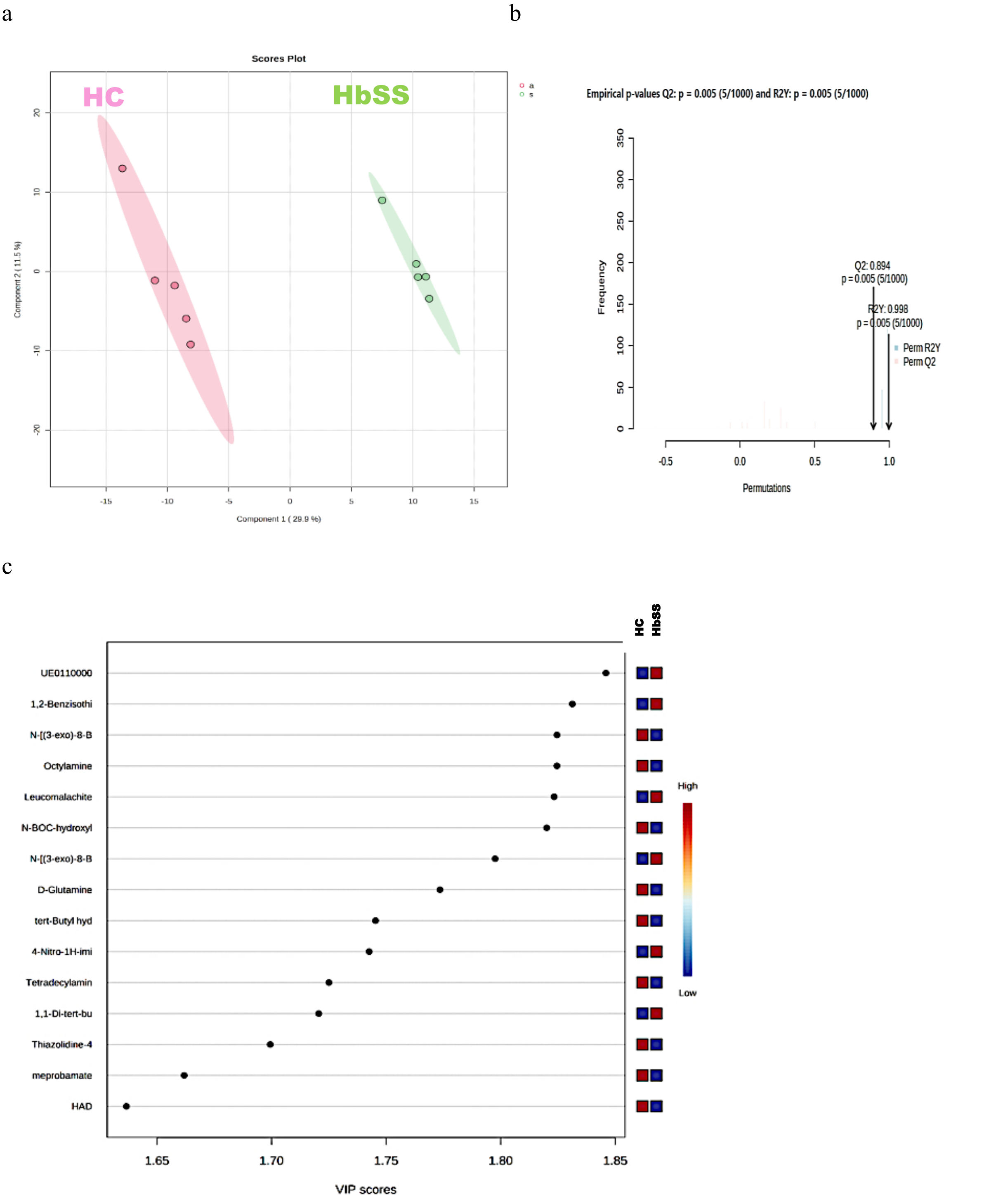


Figure S2. RBC metabolites of sickle cell disease (HbSS: SCD) and control (HC: healthy) subjects cluster separately in OPLS-DA. (a) Score plot showing separate clusters for SCD and healthy subjects. (b) Permutation test results with observed and cross-validated $R_2Y$ (fraction of variance of the X and Y matrix) and $Q_2$ coefficients (p. (c) VIP score of top-ranked important metabolites identified from OPLS-DA.

a

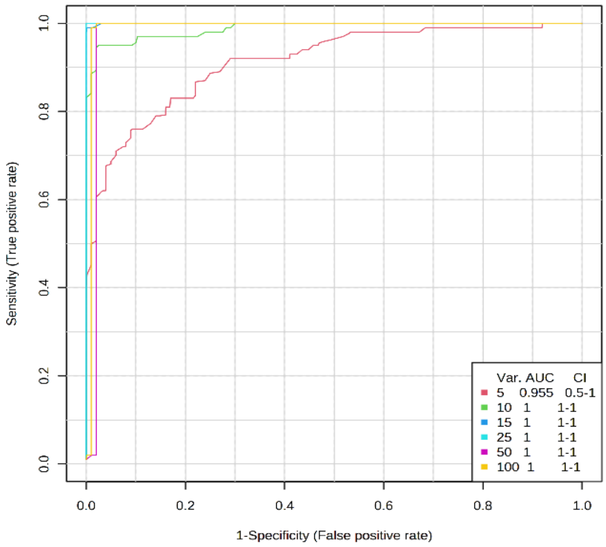


b

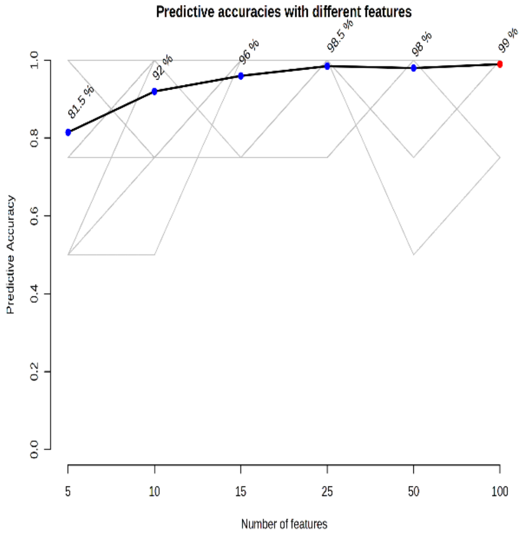


c

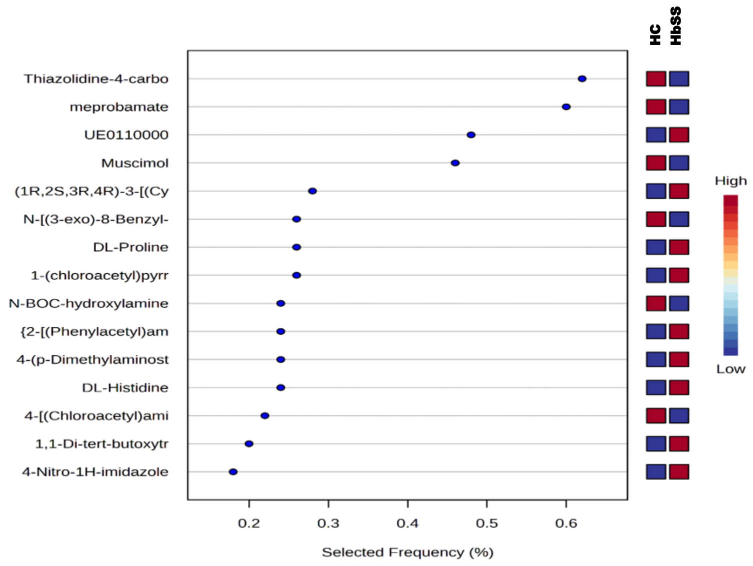


Figure S3. Model performance of six SVM classifiers for sickle cell disease(HbSS: SCD) and control (HC: healthy) subjects. (a) Each SVM classifier's ROC ( receiver operating characteristic ) curves, based on average cross-validation performance. AUCs and 95% CIs are presented in the figure legend. (b) Predictive accuracy for each SVM. The model with the highest accuracy is highlighted in red. (c) Variable importance from the SVM model with 25 metabolites. Metabolites are ranked from most to least important. The colored boxes on the right indicate metabolite concentrations in each group (HbSS: SCD and HC: healthy).

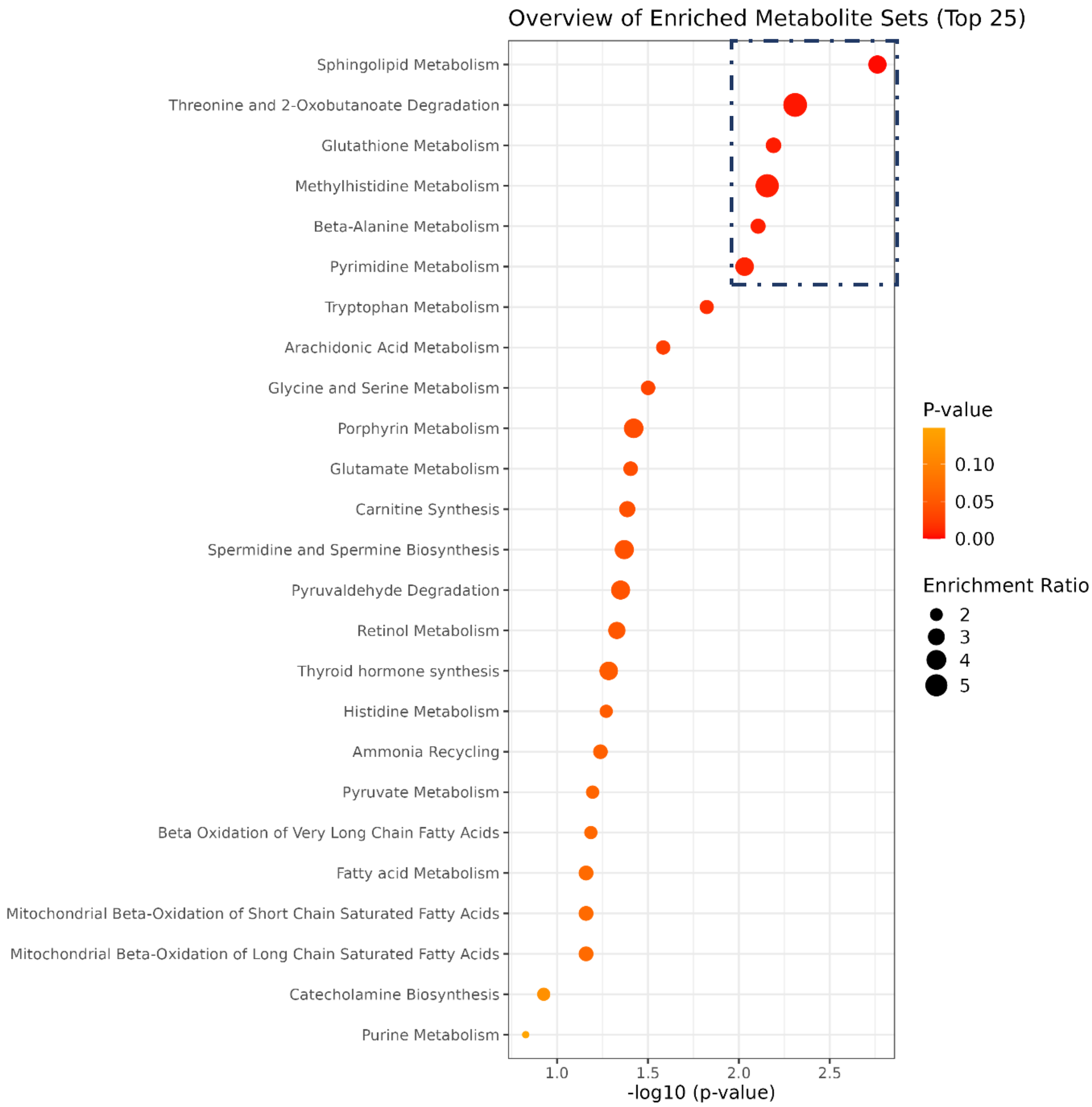


Figure S4: Top 25 enriched metabolic pathways observed in the RBC of SCD and healthy groups analyzed using KEGG database.

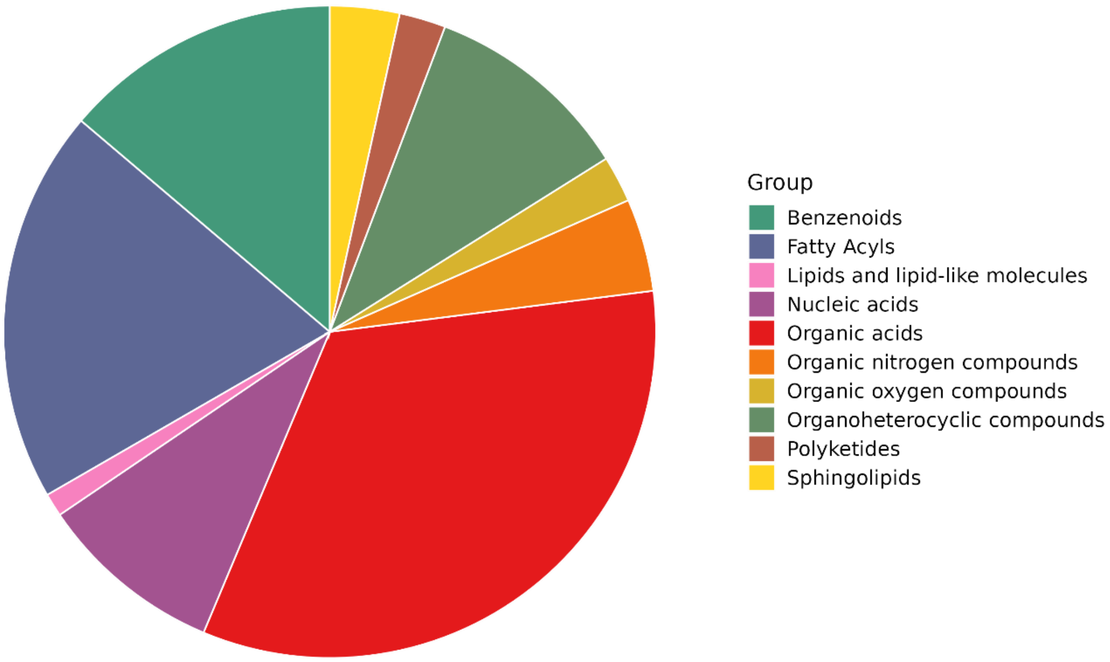


Figure S5: Identified RBC metabolites in SCD and healthy groups belong to multiple molecular classes.

a

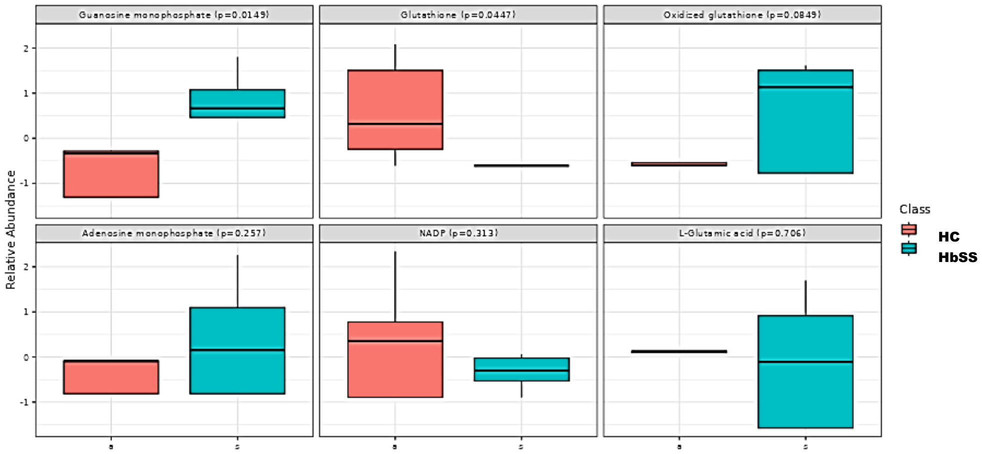


b

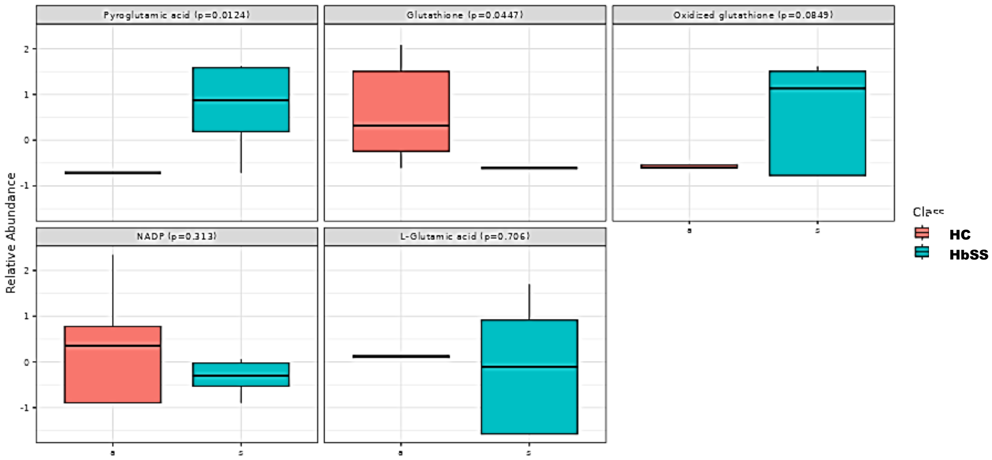


c

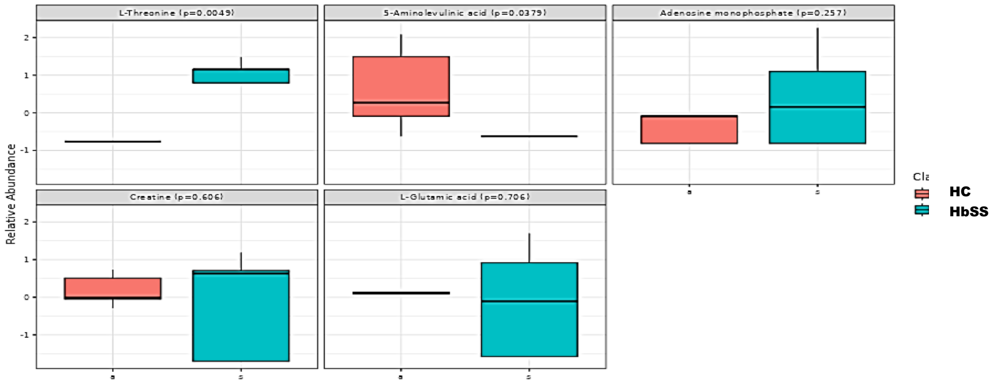


Figure S6: Relative abundance of the dysregulated metabolites presenting a) Glutamate metabolism, b) Glutathione metabolism and c) Glycine and Serine metabolism in SCD (blue) compared to the healthy (red) group, based on the KEGG analysis.
